# The dynamic motor control index as a measure of post-stroke impairments in neuromotor control

**DOI:** 10.64898/2026.04.30.26351964

**Authors:** Ashley N. Collimore-Doherty, Ruoxi Wang, David A. Sherman, Conor J. Walsh, Paolo Bonato, Terry Ellis, Louis N. Awad

**Author notes:** **Correspondence Address:** Louis N. Awad, Department of Physical Therapy, 635 Commonwealth Ave, Boston, MA 02215.

## Abstract

Measuring neuromotor control after stroke is crucial for identifying the mechanisms underlying asymmetrical walking and guiding rehabilitation. The lower extremity portion of the Fugl-Meyer (FM-LE) and the number of muscle synergies are commonly used measures, but have important limitations. The dynamic motor control index has emerged as a complementary metric, yet its relationship to established clinical measures (i.e., FM-LE), muscle synergy number, and gait biomechanics remains unclear. This study evaluated the ability of the dynamic motor control index to quantify post-stroke neuromotor impairment relative to FM-LE and muscle synergy number and examined its relationship with propulsion asymmetry. Electromyography data from 22 individuals post-stroke and 31 neurotypical controls were analyzed using non-negative matrix factorization. The dynamic motor control index and not the muscle synergy number differentiated paretic, non-paretic, and neurotypical limbs (χ2(2) = 27.57, *p* < .001). It also differed significantly between less and more impaired individuals classified by FM-LE (*p* = .05) and demonstrated good discriminative performance between these groups (AUC: 0.777, *p* = .017). The index also moderated the relationship between FM-LE and propulsion asymmetry (ΔR^2^ = 0.223, *p* = .007). These findings support the dynamic motor control index as a clinically relevant msarker of post-stroke neuromotor impairment and recovery.

## Introduction

Impaired walking after stroke is characterized by reduced speed and asymmetric stepping^1,2^, and is a major contributor to reduced quality of life^3,4^. Functional outcomes such as walking speed and endurance are commonly targeted and tracked throughout rehabilitation, while biomechanical measures of gait quality (e.g., symmetry) are often considered secondary^5^. Although both functional and biomechanical outcomes describe important aspects of walking performance, they do not directly measure impairments in neuromotor control (i.e., muscle coordination) underlying post-stroke gait. Understanding these coordination deficits is essential for personalizing rehabilitation and maximizing walking recovery. For example, individuals post-stroke may walk at similar speeds and with comparable gait quality while using different paretic limb muscle co-activation patterns^6–8^. Others compensate for key paretic limb locomotor subtasks using the non-paretic limb^9–12^, resulting in movement patterns that differ from neurotypical gait without reducing their walking speed^1,2,13^. Thus, gait impairments arise from alterations in neuromotor control that can only be addressed during rehabilitation if they are measured; functional measures of walking speed or endurance are insufficient.

The Fugl-Meyer is considered the gold-standard clinical measurement of neuromotor function after stroke^14,15^. The lower extremity portion of the Fugl-Meyer (FM-LE) evaluates reflexes, volitional movement, and coordination of the paretic limb relative to the non-paretic limb, during tasks completed in standing, sitting, or supine, with a maximum score of 34^15^. The FM-LE has been used to identify motor improvements following intervention^16–19^ and is related to community walking activity^20,21^. However, the relationship between FM-LE and neuromotor function measured by gold-standard biomechanical techniques, such as propulsion asymmetry, is unclear^22,23^, highlighting limitations in its ability to quantify and track biomechanical impairments. The FM-LE also has a well-known ceiling effect^15,24^, limiting its ability to effectively detect changes in neuromotor function, particularly among higher-functioning individuals. These limitations of the FM-LE indicate that complementary measures that more directly and sensitively quantify neuromotor control within and across limbs are needed.

It is theorized that during functional movements, the nervous system coordinates units of multiple muscles, known as muscle synergies, rather than controlling individual muscles independently^25,26^. Muscle synergies are extracted using non-negative matrix factorization (NNMF) from multi-muscle electromyography (EMG) recordings^27,28^. Neurotypical walking is typically characterized by three to four muscle synergies associated with major gait subtasks^7,8,29–32^, though reported values range from two to seven^7,22,31,30,32–35^. A reduced number of synergies is used to quantify neuromotor impairment (i.e., increased co-activation)^7,22,32^, and post-stroke, there is often a reduction in the number of synergies on the paretic limb compared to both the non-paretic limb and age-matched neurotypical controls^7,22,31,36^. Fewer synergies on the paretic limb are associated with worse propulsion symmetry^7,22^, walking speed^7,22,37^, step length asymmetry^7^, joint kinematics^37^, and clinical measures of balance^22^. However, the discrete nature and restricted range of synergy numbers limit its overall sensitivity for detecting gradations in neuromotor impairment or changes over time or following intervention^38,39^. Indeed, some people post-stroke may exhibit a similar synergy number on the paretic limb as healthy adults, despite altered muscle activation timing and marked biomechanical impairments^6,7,39,40^. Moreover, people with the same number of synergies can still display different muscle coordination patterns and levels of impairment^6,7^, limiting the ability of synergy number to distinguish between impairment severity or to capture intra- and inter-subject differences^7,39^. This limitation can be further compounded by the sensitivity of synergy number to EMG processing techniques^41,42^ and the cutoff criteria used for the NNMF algorithm^30,43^, both of which are not standardized across research groups. The wide range of synergies for neurotypical gait (i.e., 2 to 7) reflects the influence of methodological choices^42^. Together, these factors highlight the need for a more sensitive and robust measure of neuromotor control.

The dynamic motor control index for walking has recently emerged as an alternative metric of neuromotor control that measures the extent of overall muscle co-activation during walking^44,45^. Calculated by scaling the single-synergy solution derived from an individual to a neurotypical reference group^45,46^, this continuous metric enables easy comparison within and across individuals^31^. Importantly, it is less affected by EMG processing and factorization cutoff criteria than the number of synergies^42^, thus addressing key methodological limitations of synergy number-based approaches. Originally developed to characterize impaired neuromotor control in children with cerebral palsy^45^, the metric has since shown success in identifying reduced muscle coordination associated with aging^47,48^, traumatic brain injury^49^, and haemophilic arthropathy^50^, including being sensitive to the detection of impairments that are not captured by the number of muscle synergies^47,50^. More recently, the dynamic motor control index has also demonstrated enhanced clinical relevance for individuals post-stroke compared to the number of muscle synergies, with reported correlations with functional measures of balance performance, gait speed, and FM-LE scores^31^. These results suggest the dynamic motor control index may provide a more sensitive and meaningful measure of post-stroke neuromotor control, particularly for identifying biomechanically relevant impairments among individuals with the same number of muscle synergies.

The objective of this study is to replicate and extend prior work^31^ evaluating the dynamic motor control index for measuring stroke-related impairment in neuromotor control. First, we compare the ability of the dynamic motor control index versus the number of muscle synergies to differentiate between paretic, non-paretic, and neurotypical limbs. We hypothesize that the dynamic motor control index will better distinguish between limbs compared to the number of muscle synergies. Second, we assess the relationship between the dynamic motor control index of the paretic limb and the gold-standard clinical measure of post-stroke neuromotor function, the FM-LE. We examine this relationship across the full score range and also by subgrouping participants using an established FM-LE cutoff for hemiparetic impairment severity^51^. We hypothesize that the index will be related to the FM-LE, with higher dynamic motor control indices (i.e., better control) among individuals with higher FM-LE scores, and will accurately identify hemiparetic severity groups. Lastly, we evaluate the relationships between the FM-LE, the dynamic motor control index, and biomechanical gait quality, as measured by propulsion asymmetry. We hypothesize that the dynamic motor control index will be more related to propulsion asymmetry compared to the FM-LE. Together, these objectives aim to clarify the relationship between the dynamic motor control index and commonly used neuromotor, clinical, and biomechanical measures of post-stroke walking.

## Methods

### Participants

Twenty-two individuals aged 18 - 75 years in the chronic phase of stroke who could walk independently were recruited. Participants were allowed to wear assistive devices providing medio-lateral ankle support but not restricting plantarflexion; individuals requiring plantarflexion-restricting devices (solid ankle-foot orthoses) were excluded. Additional exclusion criteria included subcortical stroke, severe aphasia impairing communication (NIHSS question 1b > 1 and question 1c > 0), pain limiting walking, neglect, hemianopia, unexplained dizziness in the previous 6 months, resting heart rate outside 50 - 100 bpm, and resting blood pressure outside 90/60 - 200/110 mmHg. Data were collected as part of a larger study requiring a neutral ankle angle for additional assessments; individuals unable to achieve a neutral ankle angle were excluded. The study was approved by the Boston University Institutional Review Board. All participants provided written informed consent.

An open-source dataset with 31 neurotypical adults (45 ± 21 years) served as the control group^46^. Young (20 - 35 years, N = 18) and young-old (60 - 74 years, N = 13) groups from our prior work were combined, as no differences in dynamic motor control indices were observed among adults under 75 years^47^; data from individuals ≥ 75 years were excluded. Data collection and processing have been previously described^46,47^.

### Data Collection

Participants walked on an instrumented, dual-belt treadmill (Bertec, Columbus, Ohio, USA) for ≥ 90 seconds at their fastest safe speed. Speed was initially determined from an overground 10-meter walk test and adjusted based on participant and physical therapist perceived safety. Participants wore an overhead safety harness without body weight support and could use a handrail if needed.

During treadmill walking, bilateral EMG was recorded from 11 lower-limb muscles^30,52^ (Figure 1) following SENIAM placement guidelines^53,54^: vastus medialis, rectus femoris, vastus lateralis, soleus, medial gastrocnemius, peroneus longus, tibialis anterior, biceps femoris, medial hamstrings, gluteus maximus, and gluteus medius. Hamstring and glute muscles were collected at 1256 Hz (Avanti, Delsys Inc., Natick, MA, USA) and the rest at 1111 Hz (Quattro, Delsys Inc.)^52^. Ground reaction forces were simultaneously recorded for each leg at 2000 Hz.

**Figure 1.**
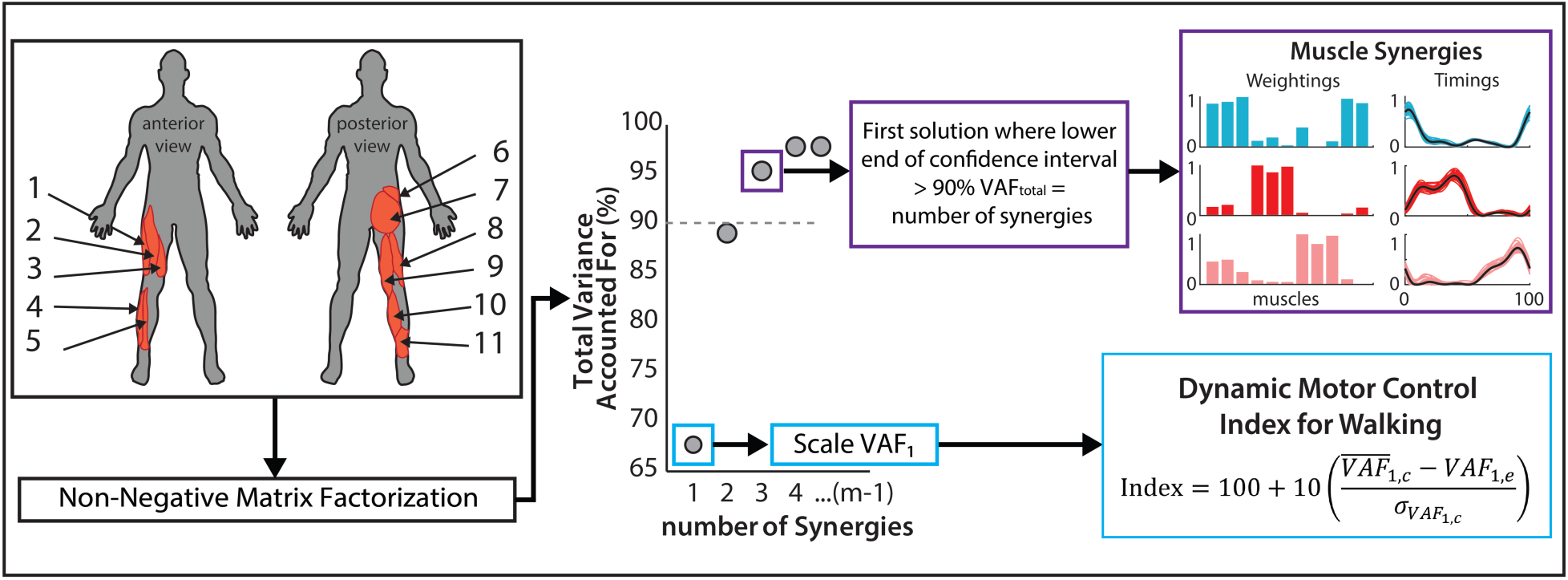
– Overview of Muscle Collection and Metric Calculations. Electromyography activity was collected from 11 muscles bilaterally: 1) Vastus Lateralis, 2) Rectus Femoris, 3) Vastus Medialis, 4) Peroneus Longus, 5) Tibialis Anterior, 6) Gluteus Medius, 7) Gluteus Maximus, 8) Biceps Femoris, 9) Medial Hamstrings, 10) Medial Gastrocnemius, and 11) Soleus. Non-negative matrix factorization was used to calculate both the number of muscle synergies and the dynamic motor control index. Abbreviations: VAF_total_: total variance accounted for; VAF_1_: variance accounted for by the one muscle synergy solution. c: neurotypical control group. e: each individual in the experimental groups.

The lower-extremity Fugl-Meyer Assessment (FM-LE) was conducted by a trained research physical therapist (maximum score = 34). Participants were stratified into more impaired (FM-LE < 21) and less impaired (FM-LE ≥ 21), using an established FM-LE, which was validated using clinical tests of endurance, speed, and balance^51^.

### Data Processing

EMG signals were high-pass filtered at 40 Hz (4^th^ order Butterworth), demeaned, rectified, low-pass filtered at 4 Hz (4^th^ order Butterworth), and resampled to 1000 Hz. EMG channels were visually inspected by two investigators, and those with poor signal quality (e.g., persistent artifact or no muscle modulation) were removed on a participant- and limb-specific basis^55^. Individual strides containing signal artifacts were also excluded, and the last 30 clean strides were used for analyses. Each stride was resampled to 101 points to normalize to the gait cycle.

Muscle synergy number and dynamic motor control index values were calculated using NNMF using a modified open-source MATLAB script^33^. To determine the number of synergies, a cutoff-criteria was set to the lower end of the confidence interval for the total variance accounted for (VAF) surpassing 90% or until the addition of a new synergy did not increase the total VAF by more than 5%^29,56^ (Figure 1, purple). The dynamic motor control index was calculated from the VAF of the one-synergy solution (VAF_1_) using the equation^45^ (Figure 1, blue):

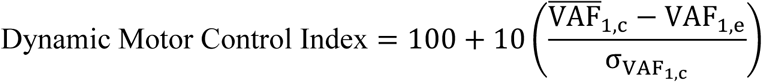

where 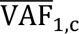 and σ_VAF_1,c__ represent the mean and standard deviation of VAF_1_ from the neurotypical control group, and VAF_1,e_ is the value for each individual in the experimental groups (neurotypical controls, non-paretic limbs, and paretic limbs).

Ground reaction forces were low-pass filtered at 15 Hz (4^th^ order Butterworth). Initial contact and toe off events were identified using a 30 N threshold. Strides were visually assessed, and the last 30 clean strides were used for analysis. Peak propulsion was calculated as the peak anterior ground reaction force during stance phase. Peak propulsion asymmetry was calculated on a step-by-step basis as the ratio of the paretic limb to the sum of both limbs, where 50% represents perfect symmetry; values < 50% indicate non-paretic leaning and > 50% indicate paretic leaning^57–59^. Variables were averaged across strides. All processing was done using Visual 3D (version 2023, HAS-Motion Inc., Kingston, ON, Canada).

### Statistical Analyses

Due to its discrete nature, differences in the number of synergies across paretic, non-paretic, and neurotypical limbs were assessed using a chi-square test, whereas differences in the dynamic motor control indices across limbs were evaluated using one-way ANOVA with Šidák-corrected post-hoc comparisons. Multinomial logistic regression was used to determine whether the number of muscle synergies or the dynamic motor control index better identified limb type, with the neurotypical limbs as the reference. The first block contained the number of muscle synergies and the dynamic motor control index, both mean-centered, and the second block included the interaction between the two metrics.

To address the second aim, Pearson correlation was used to assess the relationship between the paretic-limb dynamic motor control index and FM-LE^31,60^. An independent t-test compared paretic-limb dynamic motor control indices between individuals with less and more impairment based on the clinically validated FM-LE cutoff of 21^51^. Receiver operating characteristic (ROC) analyses further evaluated the clinical relevance and the discriminative performance of the paretic-limb dynamic motor control index with respect to impairment severity; the area under the curve (AUC) value was calculated to reveal its ability to distinguish between the FM-LE impairment groups.

Multiple linear regression was used to evaluate the relationship between the paretic-limb dynamic motor control index and propulsion asymmetry, and if the index was better related than FM-LE. The dynamic motor control index and FM-LE scores were mean-centered and added as the first block. The interaction between the two was included as a second block.

Correlation and ROC analyses were computed in MATLAB R2024b; all other analyses were computed in SPSS (Version 29.0.1.0, IBM Corp, Armonk, NY, USA). Significance was set at *p* < .05. Dynamic motor control index values were checked for normality and outliers prior to analyses.

## Results

Twenty-two individuals post-stroke (60 ± 8 years old, chronicity 6 ± 4 years, 16 males, 10 right paretic) completed the study. Non-paretic limb muscle activity was not available for one participant. Participants walked on the treadmill at an average of 0.94 ± 0.30 m/s, with a range from 0.45 to 1.36 m/s (Table 1). Eight participants have FM-LE scores below 21 and are considered to have low motor function. After inspection of each EMG channel (see *Methods*), an average of 10 ± 1 channels are available for the paretic limb and the non-paretic limb, and 11 channels for the neurotypical controls from the open-source data set^46^.

**Table 1.** Participant baseline characteristics.

| Characteristics | Mean ± SD |
| --- | --- |
| Age (y) | 59.7 ± 8.0 |
| Chronicity (y) | 6.0 ± 4.1 |
| Sex (M/F) | 16/6 |
| Paretic Side (R/L) | 10/12 |
| Fugl-Meyer Score | 21.4 ± 6.4 |
| Treadmill Speed (m/s) | 0.94 ± 0.31 |

*Note:* Four participants used an ankle brace or aircast for medio-lateral ankle support

Whereas the number of synergies (χ^2^(4) = 5.155 *p* = .272) did not differ between neurotypical controls (3.03 ± 0.40), non-paretic (3.00 ± 0.44) and paretic limbs (2.77 ± 0.42; Figure 2A), the dynamic motor control indices differed significantly across limbs (*p* < .001) and post-hoc analyses revealed significant differences between neurotypical (100 ± 10) and non-paretic limbs (88.63 ± 11.34; *p* = .025), neurotypical and paretic limbs (77.97 ± 16.52; *p* < .001), and non-paretic and paretic limbs (*p* = .008; Figure 2B).

**Figure 2.**
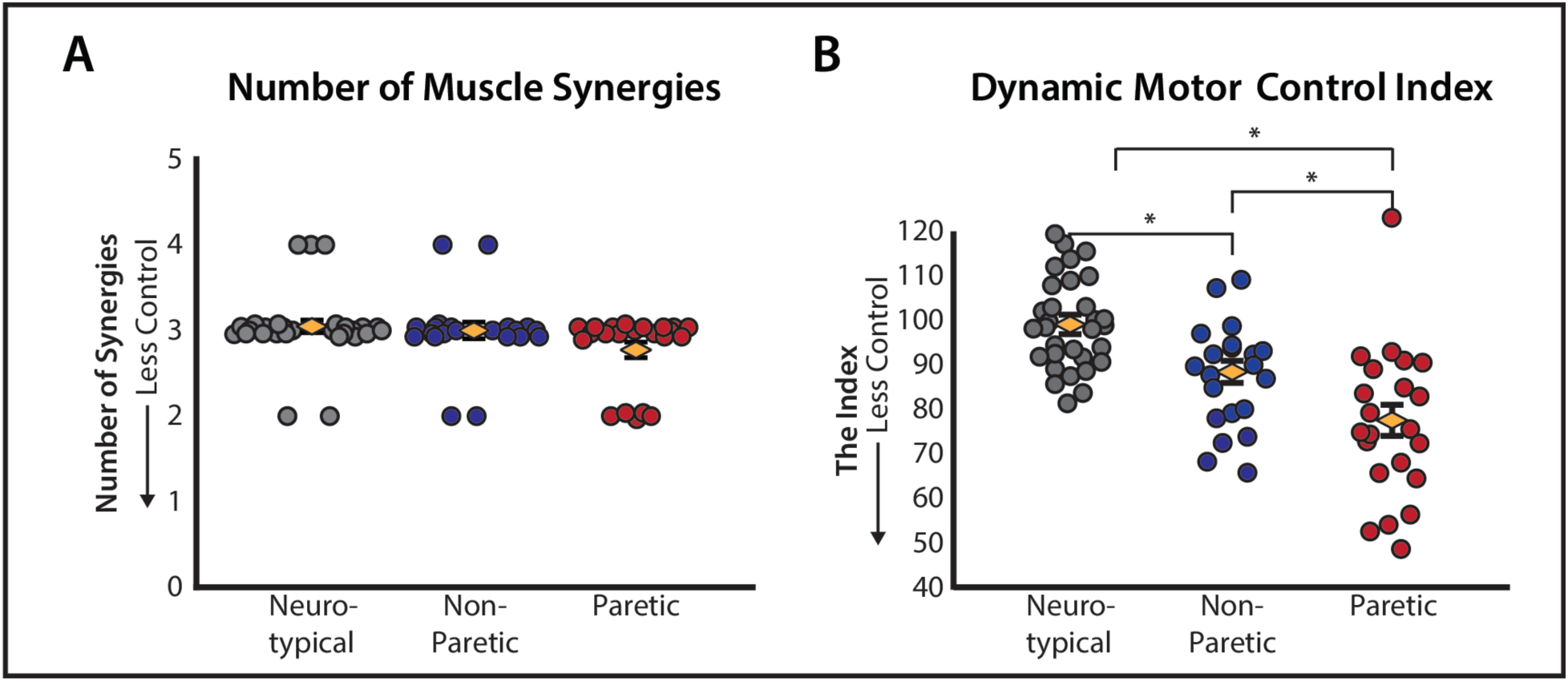
– Dynamic Motor Control Index vs. Number of Muscle Synergies. **(A)** The number of muscle synergies was not significantly different between neurotypical, non-paretic, and paretic limbs. Due to its discrete nature, jittered points are used to visualize individual synergy numbers. The averages are indicated by yellow diamond-shaped markers and error bars are standard error. **(B)** The dynamic motor control index is significantly different across all three types of limbs. The averages are indicated by yellow diamond-shaped markers and error bars are standard error.

The multinomial logistic regression model predicting neurotypical, non-paretic, and paretic limb types from the number of muscle synergies and the dynamic motor control index was significant (χ^2^ (4) = 32.83, *p* < .001, Nagelkerke R^2^ = 0.405). The dynamic motor control index was a significant predictor (χ^2^(2) = 27.57, *p* < .001), while the number of muscle synergies was not (χ^2^(2) = 0.771, *p* = .680) (Table 2). After adjusting for the number of muscle synergies, the dynamic motor control index was a significant predictor of both non-paretic (Wald χ^2^ = 8.48, *p* = .004, OR = 0.912) and paretic (Wald χ^2^ = 15.97, *p* < .001, OR = 0.863) limbs, compared to the neurotypical limbs. The interaction between the number of synergies and dynamic motor control index was not significant (*p* = .607) (Table 3).

**Table 2.**
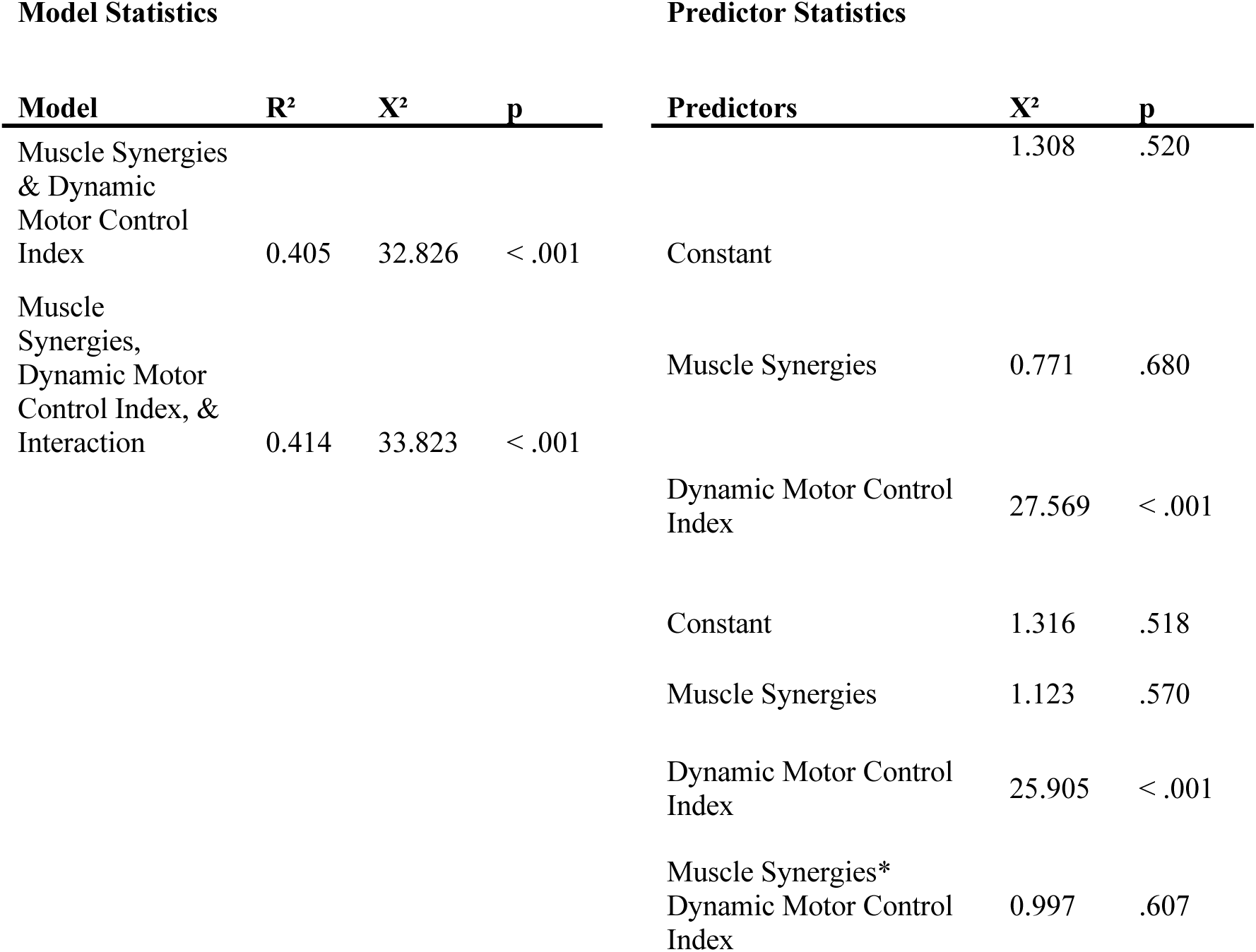
Multinomial logistic regression results: neurotypical, non-paretic, and paretic limbs.

| Model Statistics |  |  |  | Predictor Statistics |  |  |
| --- | --- | --- | --- | --- | --- | --- |
| Model | R<sup>2</sup> | X<sup>2</sup> | p | Predictors | X<sup>2</sup> | p |
| Muscle Synergies<br>& Dynamic<br>Motor Control<br>Index | 0.405 | 32.826 | < .001 |  | 1.308 | .520 |
|  |  |  |  | Constant |  |  |
| Muscle<br>Synergies,<br>Dynamic Motor<br>Control Index, &<br>Interaction | 0.414 | 33.823 | < .001 | Muscle Synergies | 0.771 | .680 |
|  |  |  |  | Dynamic Motor Control<br>Index | 27.569 | < .001 |
|  |  |  |  | Constant | 1.316 | .518 |
|  |  |  |  | Muscle Synergies | 1.123 | .570 |
|  |  |  |  | Dynamic Motor Control<br>Index | 25.905 | < .001 |
|  |  |  |  | Muscle Synergies*<br>Dynamic Motor Control<br>Index | 0.997 | .607 |

**Table 3.**
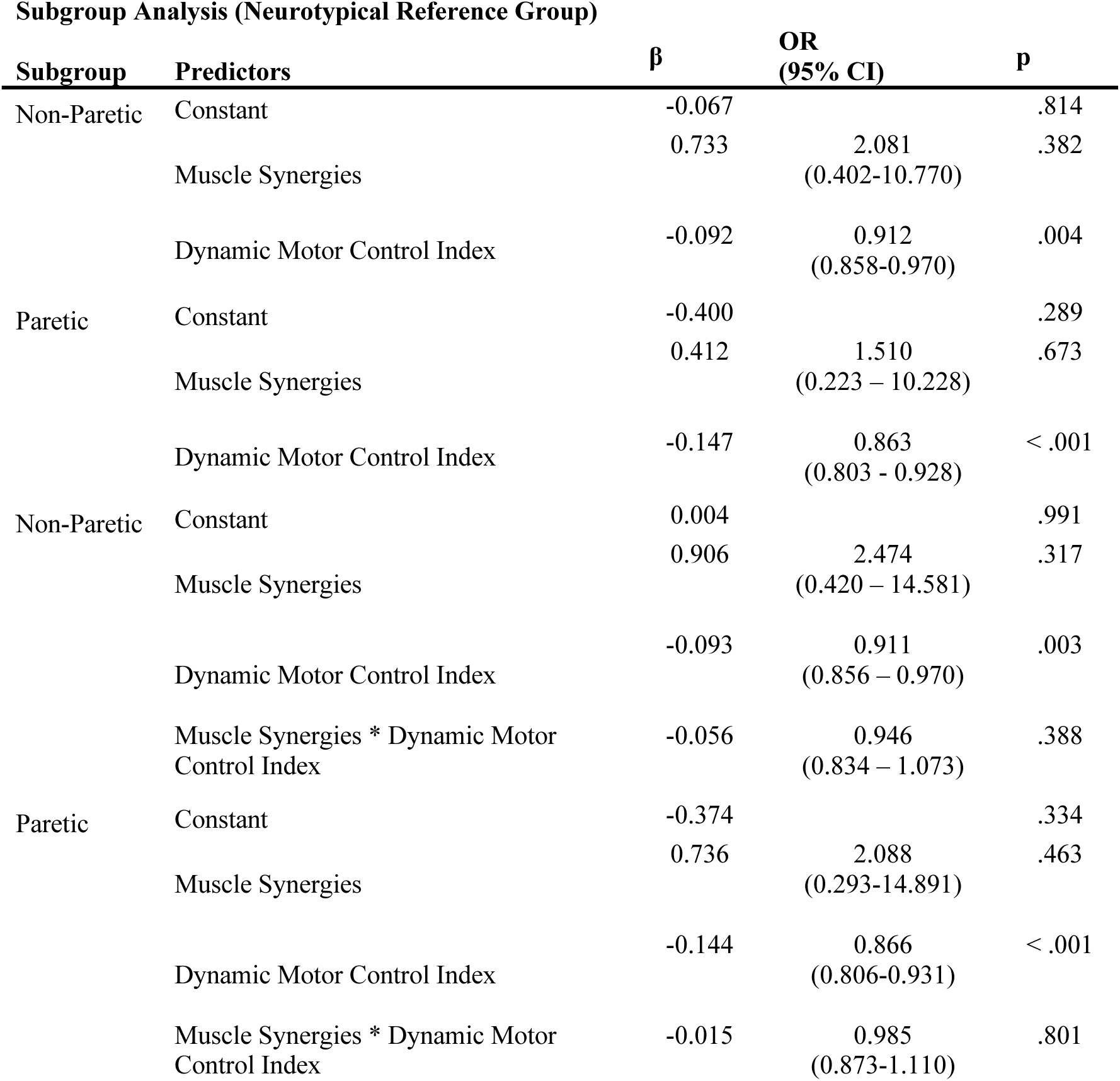
Multinomial logistic regression results: neurotypical, non-paretic, and paretic limbs (after adjusting for the number of muscle synergies)

Although only a non-significant, weak positive trend was observed between paretic-limb dynamic motor control index and FM-LE (r = 0.33, *p* = .130) (Figure 3A), the categorical split by the established FM-LE cutoff of 21 points^51^ revealed a significant subgroup difference in paretic dynamic motor control index between the more impaired (68.72 ± 12.41) and less impaired (83.25 ± 16.25) neuromotor function groups (*p* = 0.050; Figure 3B). In addition, ROC analyses showed that the dynamic motor control index could distinguish between these subgroups with high sensitivity and specificity (Figure 3C; AUC = 0.777, *p* = .017, sensitivity = 0.857, specificity = 0.750, cutoff = 74.02).

**Figure 3.**
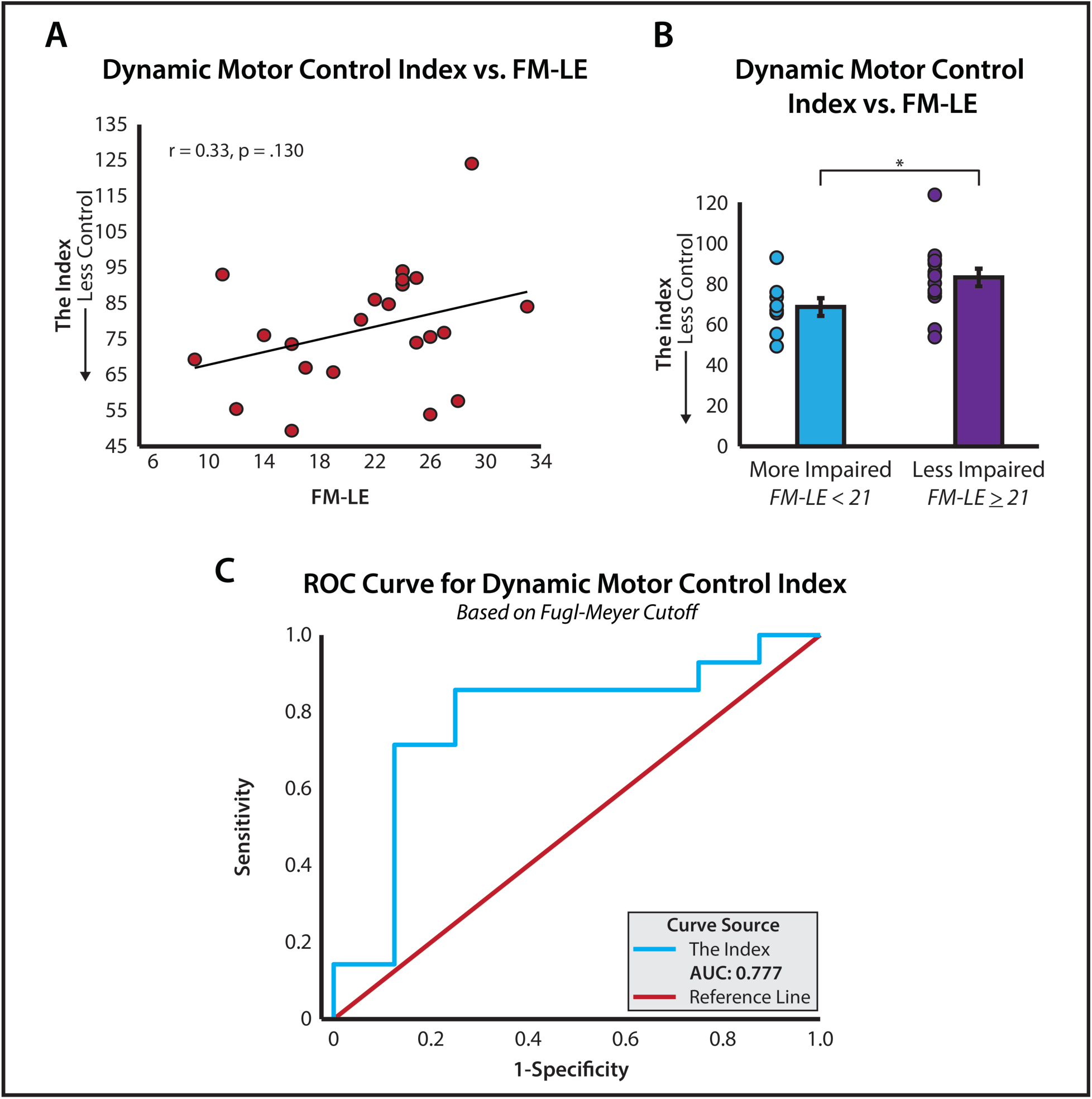
– Dynamic Motor Control Index vs. FM-LE. **(A)** There was not a significant correlation between FM-LE scores and the dynamic motor control index of paretic limbs. **(B)** When classified based on FM-LE cutoff of 21, the average dynamic motor control index of individuals with more motor impairment (FM-LE < 21) was significantly lower than those with less motor impairment. **(C)** Analysis of the ROC curve shows the dynamic motor control index could distinguish between less and more impaired individuals based on the clinical FM-LE cutoff of 21 points with high sensitivity and specificity. Abbreviations: FM-LE: the lower extremity portion of the Fugl-Meyer; ROC: receiver operating characteristic; AUC: area under the curve.

The multiple linear regression model predicting propulsion asymmetry from FM-LE and the dynamic motor control index was significant (F = 5.178, *p* = .016, R^2^ = 0.353). FM-LE was a significant predictor (*p* = .007), but the dynamic motor control index was not (*p* = .952). There was a significant change in R^2^ with the addition of the interaction between FM-LE and the dynamic motor control index (ΔR^2^ = 0.223, *p* = .007, Model R^2^ = 0.505, Model *p* = .001). Both FM-LE (*p* < .001) and the interaction term (*p* = .007) were significant predictors (Figure 4; Table 4).

**Figure 4.**
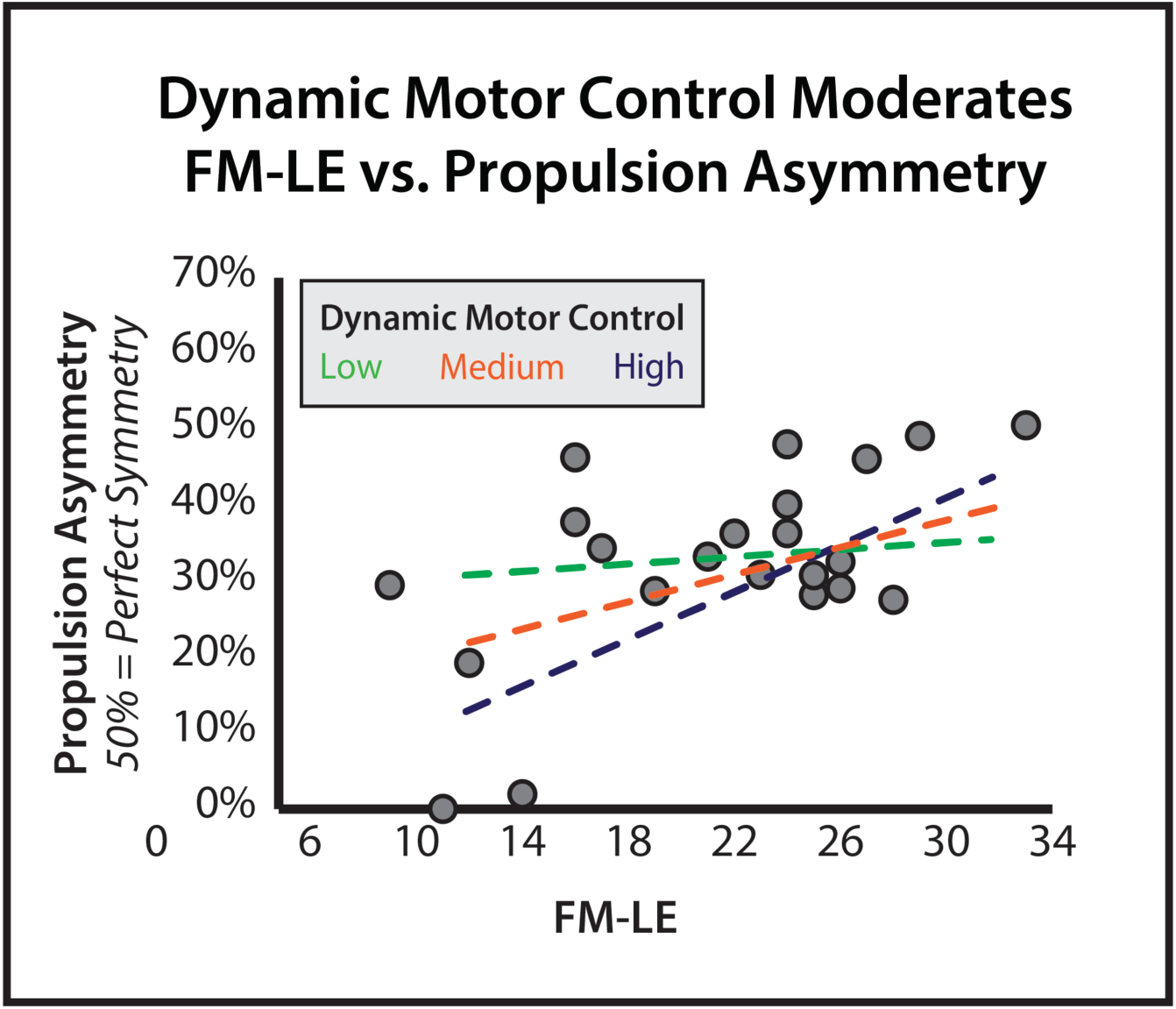
– Propulsion Asymmetry Moderation. The relationship between FM-LE and propulsion asymmetry is moderated by the dynamic motor control index of the paretic limbs. There was a strong, positive relationship between FM-LE and propulsion among individuals with high dynamic motor control indices, but no relationship (i.e., slope ∼ 0) in individuals with low neuromotor control. Abbreviations: FM-LE: the lower extremity portion of the Fugl-Meyer.

**Table 4.** Multiple Linear Regression: Predicting Propulsion Asymmetry from FM-LE and Dynamic Motor Control Index.

| Model Statistics |  |  |  |  | Predictor Statistics |  |  |
| --- | --- | --- | --- | --- | --- | --- | --- |
| Model | R<sup>2</sup> | Adj.<br>R<sup>2</sup> | F | p | Predictors | β | p |
| Paretic Dynamic Motor Control & FM-LE | 0.353 | 0.285 | 5.178 | 0.016 | Constant |  | <0.001 |
| Paretic Dynamic Motor Control, FM-LE, & Interaction | 0.576 | 0.505 | 8.140 | 0.001 | Paretic Dynamic Motor Control | -0.012 | 0.952 |
|  |  |  |  |  | FM-LE | 0.598 | 0.007 |
|  |  |  |  |  | Constant |  | <0.001 |
|  |  |  |  |  | Paretic Dynamic Motor Control | -0.178 | 0.313 |
|  |  |  |  |  | FM-LE | 0.674 | <0.001 |
|  |  |  |  |  | Interaction | 0.498 | 0.007 |

## Discussion

This study evaluated the dynamic motor control index as a measure of post-stroke impairment in neuromotor control, including its ability to differentiate limb types relative to the number of muscle synergies, its relationship with the gold-standard measure of post-stroke motor impairment (i.e., FM-LE) and impairment severity, and its relevance to biomechanical gait quality. The findings indicate that the dynamic motor control index captures clinically and biomechanically meaningful aspects of post-stroke neuromotor impairment. Specifically, dynamic motor control index outperformed the number of muscle synergies in differentiating neuromotor control across neurotypical, non-paretic, and paretic limbs, and classified impairment severity defined by the FM-LE. It also significantly moderated the relationship between propulsion asymmetry and FM-LE. These results support the dynamic motor control index as a meaningful metric of post-stroke neuromotor control impairment.

Building on prior work in multiple populations^31,47,50^, this study demonstrated that the dynamic motor control index outperformed the number of muscle synergies in identifying impairments in neuromotor control across neurotypical, non-paretic, and paretic limbs. In agreement with recent work^31^, our results showed differences in neuromotor control between the neurotypical and non-paretic limbs, which have not been previously detected using synergy number^7^. This is fundamental for the future of post-stroke rehabilitation; the non-paretic limb is often used as the reference for the paretic limb’s progress (e.g., in FM-LE) and not individually targeted by rehabilitation paradigms, despite potential deficits relative to neurotypical function. Here, the dynamic motor control index detects neuromotor impairments in both paretic and non-paretic limbs that other measures either failed to detect (synergy number) or were not designed to assess (FM-LE). Moreover, we also found significant differences in dynamic motor control indices between non-paretic and paretic limbs. This differs from the recent work exploring post-stroke dynamic motor control at comfortable walking speeds^31^, but may be related to their finding that divergence in the dynamic motor control index between limbs may increase as speed deviates from comfortable pace. Using a participant’s fastest safe speed may have enhanced the index’s sensitivity to limb-specific neuromotor control deficits that would be less apparent at comfortable speeds. These findings suggest that the dynamic motor control index may be sensitive to not only stroke-related impairments and limb-specific deficits, but also task demands such as walking speed, similar biomechanics outcomes like propulsion asymmetry^11,58,59^. Accordingly, future interventional studies using the index to track neuromotor control changes may benefit from incorporating walking conditions beyond comfortable speed.

In contrast to the dynamic motor control index, we did not observe differences in the number of muscle synergies between neurotypical and paretic limbs^7^. This result aligns with prior findings in relatively high-functioning post-stroke individuals (fast walking speeds exceeding 0.9 m/s)^31^, but contrasts with studies that included individuals with greater impairment^7^. This suggests that differences in synergy number may be more likely to emerge in lower-functioning individuals whose gait deviates more substantially from neurotypical walking. Notably, despite our relatively high-functioning cohort, the dynamic motor control index detected clear limb-specific differences, indicating higher resolution than synergy number for identifying neuromotor control impairments. Another explanation relates to the methodological influence on synergy number estimation: while the predominance of three synergies in neurotypical gait observed here is commonly reported^7,29,31^, other studies have reported four or more synergies^30,32,33,35^ due to differences in muscle selection, signal filtering, and factorization cutoff criteria^30,41–43,61^. In contrast, the dynamic motor control index is less sensitive to EMG processing approaches and does not rely on cutoff criteria, thereby reducing the influence of methodological choices on the resulting metric^42^. As a continuous, neurotypical-referenced measure centered around an intuitive normative value of 100, the index offers practical advantages that facilitate interpretation within and across individuals and studies^31,42,44,45^. Overall, the index provides a more robust and reproducible measure of neuromotor impairment during walking.

In comparing the dynamic motor control index with the FM-LE, our results support the hypothesis that this neurotypical-referenced metric reflects impairment in neuromotor function, with lower values corresponding to more atypical and more impaired motor control as defined by the established FM-LE cutoff of 21^51^. This finding extends prior works linking dynamic motor control index to gross motor and walking function in cerebral palsy^45^ and to post-stroke impairments in gait speed, balance, and neuromotor control^31^. However, at the individual level, lower FM-LE scores were not directly correlated with lower dynamic motor control indices, contrasting prior results suggesting a weak-to-moderate association at self-selected comfortable speeds^31^. This discrepancy may be attributable to several factors in this study, including a smaller sample size, faster treadmill speed, and the potential presence of distinct neuromotor control phenotypes. Some individuals with relatively high FM-LE scores exhibited low dynamic motor control indices, possibly reflecting task-dependent neuromuscular capacity revealed at faster walking speeds, which may not be apparent under comfortable conditions for some people^7,31^ and are difficult to capture by impairment-focused clinical scales^15,24^. On the other hand, some individuals with high dynamic motor control indices had low FM-LE scores. These findings suggest that individuals post-stroke may have differential neuromotor control capacities^6,7^, and that the FM-LE and dynamic motor control index may capture overlapping yet distinct aspects of neuromotor impairment. Future work should establish reliable dynamic motor control index impairment thresholds informed by task-specific clinical and biomechanics metrics, as anchoring impairment classification on FM-LE alone is limited by its ordinal scale^15^, lack of task specificity^15,22,23^, and ceiling effects^15,24^.

Lastly, inconsistent with our hypotheses, the dynamic motor control index, is not a significant indicator of propulsion asymmetry, but rather a moderator of the relationship between FM-LE and propulsion asymmetry. When multi-muscle coordination was better, FM-LE was strongly related to propulsion asymmetry (Figure 4); when coordination was poor, this relationship was absent (slope ∼ 0). This interaction suggests that FM-LE may be a reasonable surrogate for gait quality only when neuromotor control is relatively intact, which may explain previously mixed findings regarding their relationship^22,23^. These results indicate that directly associating clinical impairment scores with biomechanical gait outcomes does not comprehensively describe post-stroke gait; measuring and understanding the underlying neuromotor control provides critical insights that may influence rehabilitation strategies.

Several limitations should be acknowledged. First, this study examined a relatively high-functioning cohort of individuals post-stroke, which may have limited our ability to observe differences in the number of muscle synergies reported in prior work, including individuals with greater impairment^7^. Nevertheless, our findings align with functionally comparable cohorts^31^ and demonstrate that the dynamic motor control index can identify stroke-related and limb-specific neuromotor control impairments even within a narrower functional range. Future work, including larger cohorts with a broader range of functional abilities, is needed to further evaluate the relationship between the dynamic motor control index and synergy-based metrics and to support data-intensive approaches for impairment classification^62^. Second, this study focused only on the number of muscle synergies as a comparator. Other synergy features, such as muscle composition and temporal patterns, were beyond the scope of this work. Prior studies show that differences in composition and timing can emerge even when synergy numbers are identical^7,39,40,63^. Examining these complementary metrics alongside the dynamic motor control index may further clarify neuromotor impairments during walking. Third, EMG data availability varied across some participants due to constraints common in clinical experiments (e.g. technical interruptions, motion artifacts, and participant tolerance), which can influence synergy number calculation^61^. After excluding low-quality channels, an average of approximately ten muscles per limb were retained from the original eleven-muscle set, exceeding the minimum of eight muscles previously shown to support dynamic motor control index-based impairment detection^52^. Nonetheless, the absence of certain muscle groups (e.g., gluteal or dorsiflexor muscles) warrants further investigation of how such omissions may affect the dynamic motor control index. Finally, neurotypical control data were drawn from a publicly available dataset ^46^, with standardized walking speeds (1.1 or 1.0 m/s) and no functional data. The control group was not directly age-matched and lacked individuals between 36 and 59 years, although 40% were older than 60 years. Prior analyses among these same participants found no age-related differences in neuromotor control^47^, suggesting the neurotypical group was likely representative for present analyses.

This study demonstrates that the dynamic motor control index identifies post-stroke impairments in the neuromotor control of walking, relates to the FM-LE, and moderates the relationship between FM-LE and propulsion asymmetry. As a continuous, neurotypical-referenced metric centered around a normative value of 100, it detects stroke-related and limb-specific neuromotor impairments in ways the number of muscle synergies cannot and facilitates clearer clinical interpretation. The differences identified between non-paretic and neurotypical limbs expand current understanding of neuromotor impairments in the non-paretic limbs and support a shift in rehabilitation paradigms to address both limbs. The dynamic motor control index shows promise as a clinically useful marker of post-stroke neuromotor impairment and recovery.

## Data Availability

Data presented in this study are available from the corresponding author upon request.

## Acknowledgements

This work was supported by the Boston University Ignition Award and the NIH (5F31HD106777-02). We thank Johanna Spangler, Kimberly Ang, and Ashlyn Aiello for their assistance with data collection. We thank our study participants who generously gave their time for this research.

## Data Statement

Data presented in this study are available from the corresponding author upon request.

